# Validating an Early Pregnancy HbA1c as the Screening Test for Gestational Diabetes Mellitus: Findings from PRISMA Pakistan Cohort

**DOI:** 10.64898/2026.06.08.26355138

**Authors:** Sabahat Naz, Nida Salman Yazdani, Zahra Hoodbhoy, Yonas Ghebremihael-Weldeselassie, Azqa Mazhar, Fyezah Jehan, Aneeta Hotwani, Imran Nisar, Romaina Iqbal

## Abstract

**Background:** Early identification of gestational diabetes mellitus (GDM) is critical to improving maternal and neonatal outcomes, particularly in resource-constrained settings where universal oral glucose tolerance testing (OGTT) is burdensome. We assessed whether early-pregnancy HbA1c alone or combined with common risk factors can predict GDM and reduce the burden of OGTT requirements in a peri-urban cohort in Karachi, Pakistan.

**Methods:** We conducted a secondary analysis of the Pregnancy Risk Infant Surveillance and Measurement Alliance (PRISMA) Pakistan cohort. Women enrolled before 20 weeks’ gestation with available early-pregnancy HbA1c and a 2-hour 75g OGTT at 24–28 weeks were included. We externally validated GDM prediction models originally developed in the STRiDE-India cohort. Model performance was evaluated using receiver operating characteristic (ROC) curves and area under the curve (AUC). We assessed four models: HbA1c alone (Model 1a); age, BMI, and family history of diabetes mellitus (FH DM) (Model 1b); HbA1c combined with age, BMI, and FH DM (Model 2); and an extended model, i.e., Model 2 combined with socioeconomic status, gestational age, parity, systolic and diastolic blood pressure (Model 3). A dual-threshold approach was applied to assess rule-in and rule-out performance.

**Results:** Among 2,489 women, GDM incidence was 7.5% (n=186). Models with a broader set of predictors demonstrated higher AUC values, with Model 2 achieving an AUC of 0.61 (95% CI: 0.57–0.66). Including additional factors (Model 3) did not further improve predictive ability (AUC: 0.62; 95% CI: 0.58–0.66). In addition, at predefined thresholds, Model 2 achieved sensitivity of 73.7% (rule-out) and specificity of 83.5% (rule-in), with potential to reduce OGTT requirements (58.5%).

**Conclusions:** Early-pregnancy risk stratification using HbA1c combined with simple clinical predictors offers a pragmatic approach to streamline GDM screening among high-risk pregnant women. A dual-threshold strategy using Model 2 could reduce reliance on universal OGTT while prioritizing high-risk women for confirmatory testing.

**What is already known on this topic:** Studies have indicated that HbA1c may be an acceptable biomarker for identifying high-risk pregnant women with GDM and hence reducing the unnecessary burden of OGTT in resource-constrained settings. However, its ability to predict Pakistani women at high risk of GDM is unclear.

**What this study adds:** Early pregnancy HbA1c, together with age, BMI, and family history of diabetes achieved an AUC of 0.61 (95% CI: 0.57–0.66), with 58.5% of OGTTs could be avoided using this method.

**How this study might affect research, practice or policy:** Early pregnancy HbA1c-based prediction model is best positioned as pragmatic triage strategies to prioritize women for OGTT rather than replacing it. Such approaches may optimize resource use, particularly in settings where access to diagnostic testing is limited.

## Background

The burden of gestational diabetes mellitus (GDM) is escalating in low-and middle-income countries (LMICs), such as Pakistan, with a recent systematic review and meta-analysis reporting an overall GDM pooled prevalence of 16.7% (1). Since GDM is associated with adverse maternal and neonatal outcomes (2), this high prevalence highlights the urgent need for timely identification and early management of GDM to mitigate these outcomes. The protocols for GDM screening and diagnosis thresholds vary across countries, with a 2-hour 75g oral glucose tolerance test (OGTT) at 24–28 weeks’ gestation being widely considered as the gold standard (3). However, its invasive, time-intensive nature and fasting requirements reduce coverage and equity (4), with studies reporting suboptimal OGTT test rates, particularly from LMICs (5–7)

Since OGTT is logistically challenging, an alternate early biomarker is being explored globally (8–10). For instance, New Zealand’s clinical guidelines recommend screening for GDM during early pregnancy using glycosylated hemoglobin (HbA1c) (11). In addition, the American Diabetes Association (ADA) “Standards of Care in Diabetes” 2024 also suggests screening pregnant women for early abnormal glucose metabolism using HbA1c (12). HbA1c is a simple, easy-to-administer test used to identify and manage people with type 2 diabetes mellitus (T2DM) (13). Unlike OGTT, HbA1c does not require fasting or multiple blood draws and reflects average glycemic control for up to 3 months, with generally lower intra-individual variability (13). Studies have indicated that HbA1c may be an acceptable biomarker for identifying high-risk pregnant women with GDM, thereby reducing the unnecessary burden of OGTT in resource-constrained settings (9, 10). In addition, the rising prevalence of GDM and variability in screening tests and diagnostic cut-offs highlighted the need for simplified, minimally invasive, or non-fasting screening pathways that can be implemented in routine antenatal care (ANC) services, particularly in high-volume LMIC primary healthcare centers (PHCs) paired with a clear referral pathway to prioritize high-risk pregnant women. However, its ability to predict Pakistani women at high risk of GDM is unclear. Therefore, we aimed to determine whether early-pregnancy HbA1c, both independently and in combination with common risk factors such as age, body mass index (BMI), and family history of diabetes mellitus (FH DM), can predict GDM and reduce the burden of OGTT requirements, in a peri-urban community-based pregnancy cohort in Karachi, Pakistan.

## Methods

### Study design and participants

The Pregnancy Risk Infant Surveillance and Measurement Alliance (PRISMA) is a multi-country, open cohort study that aims to investigate and harmonize pregnancy risk factors and their association with adverse pregnancy outcomes in LMICs (14). The PRISMA study is being conducted in five countries, including India, Kenya, Ghana, Zambia, and Pakistan. The Pakistan cohort has been described in detail previously (15). Briefly, the Pakistan site (hereafter referred to as PRISMA Pakistan) comprises two peri-urban communities (Rehri Goth and Ibrahim Hyderi) in Karachi, Pakistan, which provide a catchment area of approximately 200,000 individuals. Routine maternal and child health surveillance has been ongoing at these sites for more than a decade (16). Two PHCs at these sites, operated jointly by the Department of Pediatrics and Child Health, Aga Khan University (AKU), and VITAL Pakistan Trust (VPT) (17), provide free outpatient ANC services, including pregnancy ultrasound, and comprehensive care for children under five years of age (16). For delivery, pregnant women are registered at a midwife-led partner facility while women requiring higher-level care are referred to a public-sector tertiary care facility. Pregnant women aged 15-49 years who were residents of the study sites, had a viable pregnancy confirmed by ultrasound, and had a gestational age less than 20 weeks were eligible to participate in the PRISMA Pakistan study.

This secondary analysis used PRISMA Pakistan cohort data collected from September 22, 2022, to November 22, 2024. Among women enrolled before 20 weeks’ gestation, we included those who had an HbA1c measurement at the time of enrollment and completed a 2-hour 75g OGTT at the 24-28 week ANC visit. Women with overt diabetes, defined as Hba1c values greater than 6.5%, and those without a completed OGTT were excluded from the analysis.

### Data collection

The primary study’s data collection was facilitated by a custom-built Android application that recorded all relevant information at each ANC visit. Midwives conducted data collection at each ANC visit at the PHC. At enrolment, maternal demographics, medical and obstetric history, alongside current pregnancy information were obtained, including age, parity, family history of diabetes, and household assets. Gestational age was determined by using the ultrasound at enrolment visit, when HbA1c was also measured. Physical examination measures, including systolic and diastolic blood pressure, weight, and MUAC, were recorded at each ANC visit using standard procedures. The OGTT was conducted at the 28-week ANC visit, and all women with completed OGTT results were included in the analysis. A woman was diagnosed with GDM if one or more OGTT values were equal to or exceeded the specified glucose threshold as recommended by the International Association of Diabetes and Pregnancy Study Groups (IADPSG) (18).

### Early pregnancy HbA1c as a screening test

The screening approach includes venous HbA1c measured in early pregnancy, evaluated both as a standalone predictor and as part of a composite risk score that incorporated maternal age, BMI, and FH DM. This screening tool was originally developed and validated separately across three prospective cohorts, STRiDE-India (19), STRiDE-Kenya (20), and PRiDE-UK (21). STRiDE-India was set up in seven centers in South India, STRiDE-Kenya in seven centers in Western Kenya, and PRiDE-UK across ten centers in the UK. Further details on the STRiDE (22) and PRiDE cohorts (21) are provided elsewhere. The original models were derived using Poisson regression models where the outcome variable was the presence or absence of GDM based on the OGTT results at 24–28 weeks’ gestation. Separate models were evaluated to identify how HbA1c alone (model 1a) and in combination with other risk factors measured at the time of enrollment, such as maternal age, BMI, and Family history (FH) DM (model 1b), HbA1c adjusted for age, BMI, and FH DM (model 2), model 2 along with other variables, such as gestational age (GA), parity, socioeconomic status (SES), systolic blood pressure (SBP) and diastolic blood pressure (DBP) (model 3) predicted GDM. In the original work (9), the composite risk score was designed for early-pregnancy risk stratification rather than diagnosis, with predefined thresholds proposed to rule in or rule out GDM risk. i.e., those at higher risk (rule-in GDM), lower risk (rule-out GDM), and medium risk (requires an OGTT at 24-28 weeks), with an estimated 50% to 64% reduction in the need for OGTTs across different populations. We externally validated the GDM risk prediction models in our cohort using the equations from the STRiDE-India cohort to calculate the composite risk thresholds (Table S1). We adopted the equations from the STRiDE-India cohort because key risk attributes, such as ethnicity, sociodemographic factors, body composition, and other obstetrical factors, are similar within the South Asian population (23).

### Statistical analysis

Baseline characteristics of the women are summarized by GDM status. Frequencies and percentages are presented for categorical variables and means ± standard deviations are presented for continuous variables. Using household asset data, we derived a household wealth index via principal component analysis (PCA), excluding variables with limited variability (present in >95% or <5% of the sample). We categorized the resulting index into low, middle, and high-wealth groups to capture socioeconomic disparities.

Model performance was assessed using receiver operating characteristics (ROC) curve analysis, and discrimination was quantified by estimating the area under the ROC curve (AUC) with 95% confidence intervals. We evaluated the original model specifications, including Model 1a (HbA1c alone), Model 1b (maternal age, BMI, and FH DM), Model 2 (HbA1c combined with age, BMI, and FH DM), and Model 3 (HbA1c, age, BMI, FH DM, SES, GA, parity, SBP, and DBP).

To assess the clinical utility of the screening approach, we applied the predefined two-threshold strategy proposed in the original study (9). ROC analysis was used to identify the two threshold points: a lower threshold to achieve high sensitivity for ruling out women unlikely to develop GDM, and an upper threshold to achieve high specificity for ruling in women at high risk of developing GDM. Using the two-threshold approach, we ruled in women who were at a higher risk of GDM and eligible for early identification and management and ruled out those who were at a lower risk of GDM and unlikely to require an OGTT. Women with risk scores between the two thresholds were classified as intermediate (moderate) risk and recommended to undergo OGTT at 24–28 weeks’ gestation (refer to Table S2). Sensitivity and specificity were calculated at each threshold. The proportion of OGTTs that could potentially be avoided using this rule-in and rule-out approach was estimated for each model. All analyses were conducted using STATA (version 17).

## Results

Among 11457 pregnant women screened, 3698 provided consent and enrolled in the parent PRISMA study. After excluding women with missing data, overt diabetes and incomplete OGTT result, 2485 women were included in the final analysis (Figure 1)

**Figure 1.**
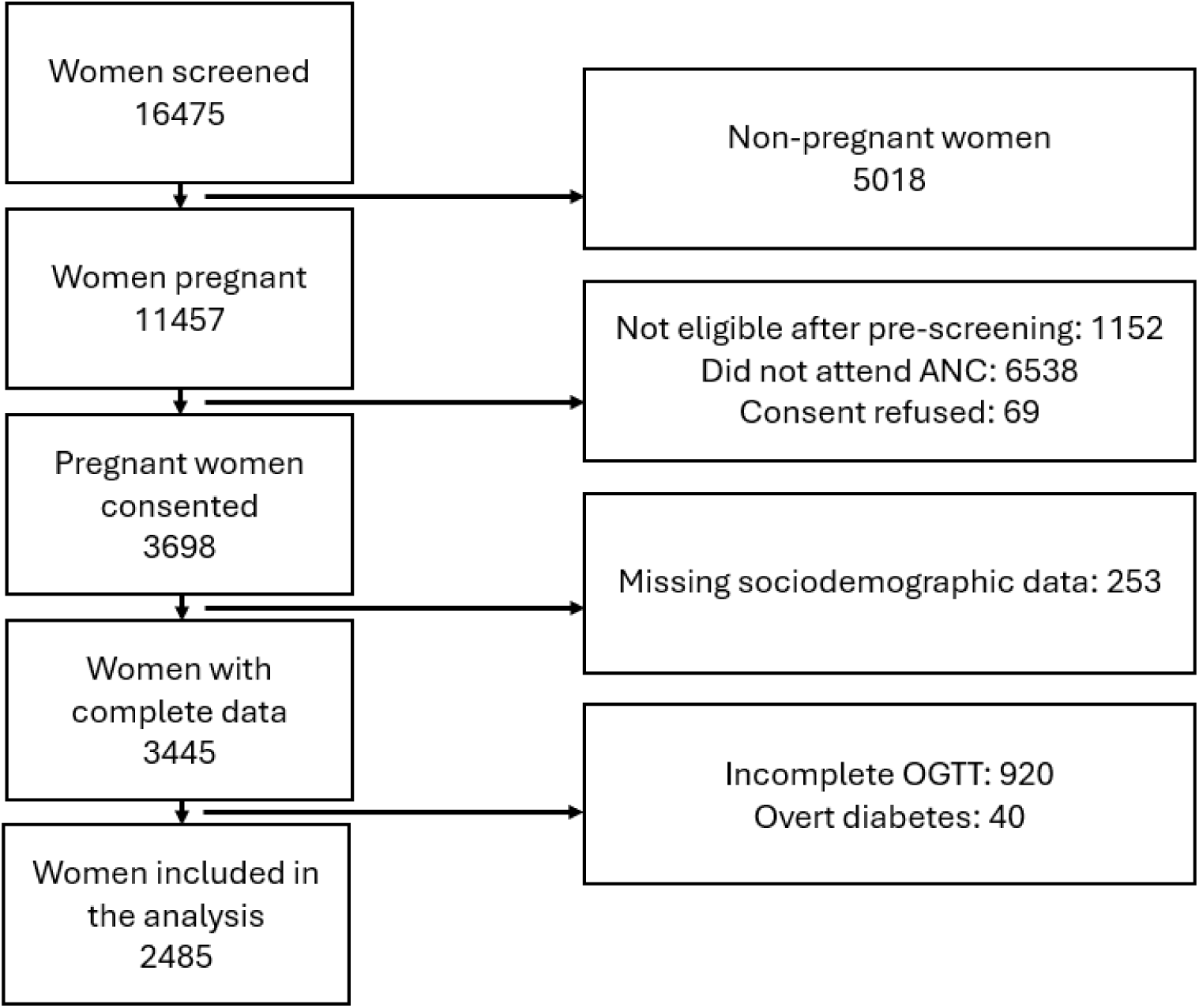
Flow diagram of PRISMA women included in the analysis.

The PRISMA Pakistan cohort had an incident rate of 7.5% (n=186) for GDM. Women with GDM were significantly older than women without GDM (28.1 ± 6.2 vs. 26.6 ± 5.9 years; p value <0.001), had higher BMI (23.6 ± 5.9 vs. 22.0 ± 4.7 kg/m2; P <0.001), and a significantly higher proportion of women with GDM (n=34, 18.3%) had a self-reported family history of diabetes than those without GDM (n=278, 12.2%). The baseline characteristics of women with and without GDM in the PRISMA cohort are presented in Table 1.

**Table 1.**
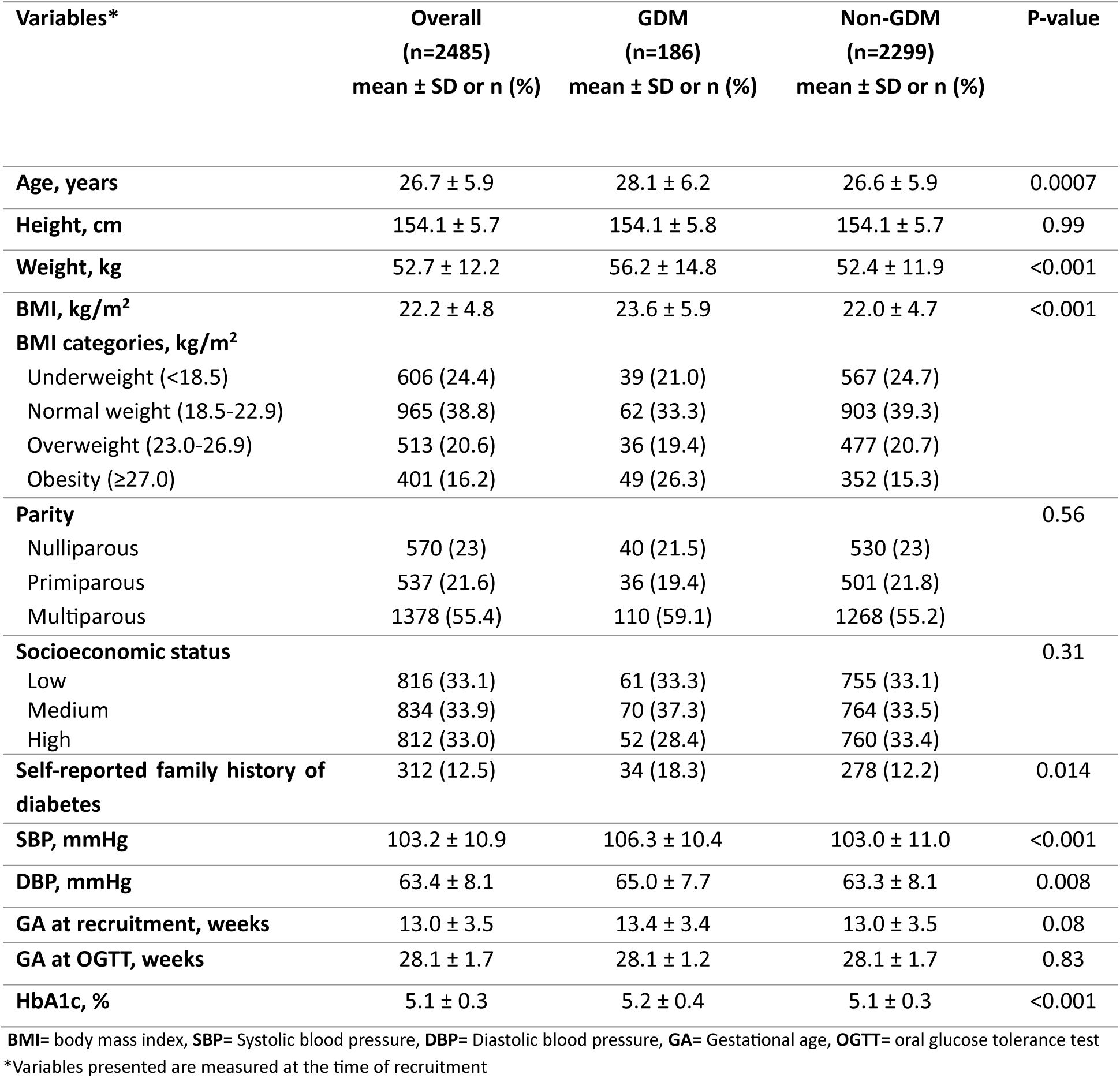
Maternal characteristics in early pregnancy in the PRISMA cohort.

ROC curves showed comparable discrimination across the models (Figure 2), particularly for Model 1a (HbA1c only) and 1b (age, BMI, FH DM), with an AUC of 0.59 (95% CI: 0.55–0.63) and 0.59 (95% CI: 0.54–0.63), respectively. Models with a broader set of predictors demonstrated higher AUC values, with Model 2 (HbA1c, age, BMI, and FH DM) achieving an AUC of 0.61 (95% CI: 0.57–0.66). However, including additional factors such as socioeconomic status, gestational age, parity, and systolic and diastolic blood pressure (Model 3) did not further improve the predictive ability (AUC: 0.62; 95% CI: 0.58–0.66).

**Figure 2.**
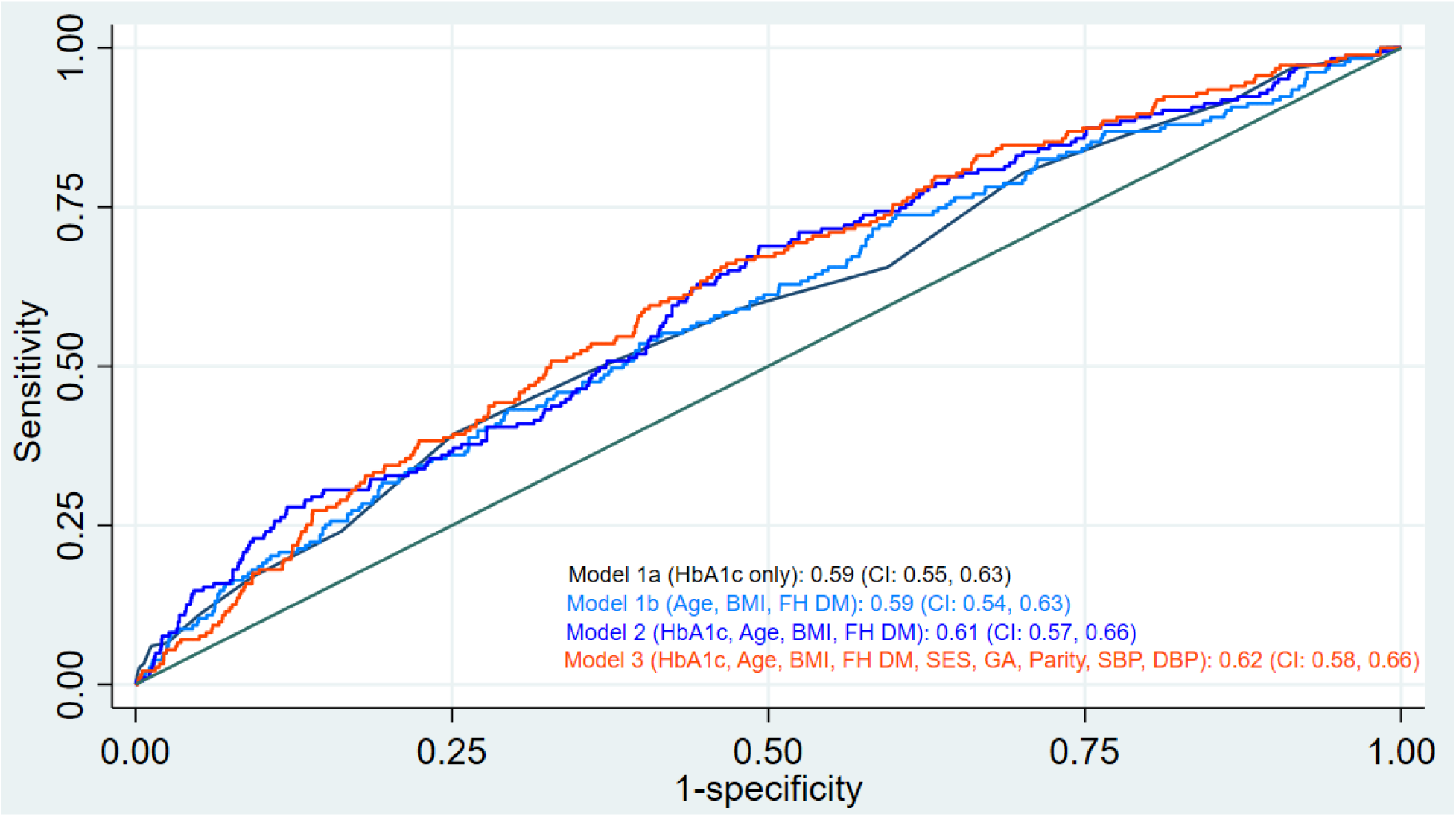
ROC curve and AUC of simple and composite prediction models.

We applied a two-threshold approach to evaluate the model’s performance for both ruling in and ruling out GDM. Threshold scores were selected based on the OGTT diagnostic cut-offs. For each model, we calculated and presented the corresponding sensitivity and specificity at the lower (rule-out) and upper (rule-in) thresholds in Table 2. The proportions of OGTT that can be avoided by this method were similar for model 1a (HbA1c alone) (47.4%) and model 1b (age, BMI, FH DM) (47.1%). The proportion of total OGTTs avoided improved to 58.5% in model 2 and 64.2% in model 3. However, Model 2 demonstrated a more balanced performance, with acceptable sensitivity (73.7%) at the rule-out threshold and acceptable specificity (83.5%) at the rule-in threshold. The detailed baseline characteristics by risk stratification for ruling in or out participants are presented in Table S2 for model 2 and Table S3 for model 3. The mean baseline age, BMI, systolic and diastolic blood pressure were higher among women ruled in with GDM (higher risk), lower among those ruled out (lower risk), and in between for those requiring an OGTT (medium risk) for both models.

**Table 2.** Proportion of OGTTs avoided using the rule-out and rule-in approach using sensitivity and specificity thresholds.

|  | Threshold scores | Sensitivity | Specificity | % of OGTTs avoided | Total % of OGTTs avoided |
| --- | --- | --- | --- | --- | --- |
| **Model 1a** |  |  |  |  | 47.4% |
| Rule-out threshold | 4.9% | 86.6% | 21.5% | 21.0% |  |
| Rule-in threshold | 5.4% | 39.8% | 74.8% | 26.4% |  |
| **Model 1b** |  |  |  |  | 47.1% |
| Rule-out threshold | 0.150 | 76.9% | 33.4% | 32.6% |  |
| Rule-in threshold | 0.224 | 22.0% | 86.1% | 14.5% |  |
| **Model 2** |  |  |  |  | 58.5% |
| Rule-out threshold | 0.164 | 73.7% | 41.8% | 40.8% |  |
| Rule-in threshold | 0.229 | 30.7% | 83.5% | 17.7% |  |
| **Model 3** |  |  |  |  | 64.2% |
| Rule-out threshold | 0.152 | 60.7% | 57.3% | 55.8% |  |
| Rule-in threshold | 0.236 | 12.6% | 92.1% | 8.4% |  |

**Model 1a** tested the performance of HbA1c alone. **Model 1b** tested the performance of age, BMI, and family history of diabetes. **Model 2** tested the performance of HbA1c, age, BMI, and family history of diabetes. **Model 3** tested the performance of HbA1c, age, BMI, and family history of diabetes, SES (wealth index), GA, parity, SBP and DBP
OGTT=oral glucose tolerance test, SES=socioeconomic status, GA= gestational age, SBP= systolic blood pressure, DBP=diastolic blood pressure
Women below the rule-out threshold will be at low risk of developing GDM (and hence will not require an OGTT); those who were above the rule-in threshold will be at high risk of developing gestational diabetes (and hence identified and managed as gestational diabetes from early pregnancy); and those who were in between will be medium risk and recommended to have an OGTT.

## Discussion

We evaluated whether an early-pregnancy HbA1c, both independently and in combination with common risk factors such as age, BMI, and FH DM, can predict GDM in a peri-urban community-based pregnancy cohort in Karachi, Pakistan. We found that early-pregnancy HbA1c was associated with subsequent GDM, with the model combining HbA1c with age, BMI, and FH DM (Model 2) appearing to offer the most clinically balanced performance, with 58.5% of OGTTs avoidable using this method. In contrast, the expanded model (Model 3) achieved a greater reduction in OGTT requirements; however, it reduced the test’s sensitivity and was therefore not considered an ideal model.

Our findings are consistent with the STRiDE-India cohort study, in which composite models combining HbA1c with age, BMI, and FH DM performed better than HbA1c alone by providing the greatest reduction in OGTT requirements (49.8%) compared to other models, with good sensitivity and specificity at the rule-out and rule-in thresholds, respectively (9). We observed the same ranking of models, with Model 2 (combining HbA1c with age, BMI, and FH DM) performing better than the HbA1c-only (Model 1a) and risk-factor-only (Model 1b) approaches. In addition, Model 2 represents the most pragmatic balance, achieving moderate sensitivity and acceptable specificity while maintaining a meaningful reduction in OGTTs (up to 58.5%). This aligns with emerging evidence suggesting that hybrid models combining biochemical markers with clinical predictors can modestly improve discrimination while maintaining feasibility in routine care, particularly in low-resource settings where implementing comprehensive testing is challenging (24, 25).

GDM screening using OGTT is both logistically challenging and financially prohibitive in resource-constrained settings like Pakistan. Therefore, developing non-invasive risk stratification tools to facilitate early identification of GDM risk is a public health priority. In this context, several risk scores have been developed, demonstrating varying discriminatory performance (26–29). For instance, Naylor et al. proposed an early model in multi-ethnic Western populations using simple predictors such as age, race, and pre-pregnancy BMI, and showed a 34.6% reduction in the OGTT requirement (26). Gao et al. developed a more comprehensive model among Chinese women, incorporating both early pregnancy clinical factors such as maternal age, BMI, height, FH DM, systolic blood pressure, and alanine aminotransferase and modifiable behavioral risks such as physical activity, sitting time at home, passive smoking, and weight gain, achieving moderate discrimination (AUC 0.71) (27). A Tanzanian model used only three easily measurable indicators (MUAC, history of stillbirth, and FH DM), with a discriminatory ability (AUC) of 0.64 (28). Kumar et al. introduced an AI-based model in Singaporean women using clinical predictors such as age, blood pressure, prior GDM, and ethnicity, with an outperformed discriminatory ability (AUC) of 0.82 (29). Overall, these tools demonstrate moderate-to-good predictive performance; however, their applicability among South Asian women is limited by differences in risk factors, such as ethnicity, sociodemographic factors, body composition, and other obstetrical factors. In addition, a lack of data on important risk factors, such as pre-pregnancy BMI, is another important barrier. This study provides one of the few external validations of early-pregnancy GDM risk-stratification models in a South Asian population, particularly using HbA1c, thereby addressing an important evidence gap. The use of a community-based cohort enhances generalizability to real-world primary care settings, where implementing universal OGTT is often challenging. In addition, the study compares multiple models, including HbA1c alone and in combination with readily available clinical predictors, enabling cross-comparison of sensitivity, specificity, and potential reductions in OGTT burden. The application of a dual-threshold (rule-in/rule-out) approach further strengthens clinical relevance by reflecting pragmatic decision-making pathways.

However, our cohort had a lower GDM prevalence (7.5%) than the pooled prevalence observed in other parts of Pakistan (16.7%) (1), which may have influenced model discrimination and predictive performance due to differences in baseline risk, population characteristics, and care-seeking patterns in this external validation setting. Therefore, these findings should be interpreted cautiously. However, it is important to note that many of these higher estimates are derived from hospital-based samples, which are more likely to include women at elevated risk and therefore may overestimate the true population-level burden. In contrast, our findings are based on a community-based sample, likely to provide a more representative estimate of GDM prevalence in real-world settings. Similarly, although early pregnancy risk stratification, particularly using combined models, can meaningfully reduce the need for universal OGTT, the low rule-in sensitivity across models reinforces that such approaches are better suited as triage tools rather than replacements for OGTT. This aligns with existing evidence indicating that first-trimester HbA1c, alone or in combination with risk factors, can identify women at elevated risk but lacks sufficient accuracy to serve as a standalone diagnostic test (9, 30).

Furthermore, BMI measurement in early pregnancy does not reflect actual pre-pregnancy status, and family history is self-reported, which may have influenced their predictive performance. However, this limitation reflects real-world conditions, particularly in marginalized communities where women often lack prior health awareness or present later to care. The modest rule-in sensitivity across models indicates a risk of missed early cases if used in isolation, limiting their role as standalone diagnostic tools. In addition, the findings are based on a specific cohort and may require further validation in diverse South Asian settings before broader implementation.

## Conclusion

Early-pregnancy GDM risk stratification using HbA1c alone or in combination with readily available clinical predictors demonstrates the potential to substantially reduce the need for universal OGTT, particularly when applied using a dual-threshold (rule-in/rule-out) approach. While more comprehensive models improved specificity and maximized reductions in OGTT requirements, all approaches showed limited rule-in sensitivity, indicating that a considerable proportion of GDM cases would be missed if a standalone tool was used. These findings suggest that such models are best positioned as pragmatic triage strategies to prioritize testing rather than replacing OGTT entirely. The next step requires implementation studies to evaluate how this model performs in real-world clinical practice, including its feasibility, acceptability, impact on screening pathways, and effectiveness in safely guiding routine care.

## Authors’ Contributions

RI conceptualized the idea. ZH, NSY, AM, AH, FJ, and IN are part of the main PRISMA study. SN was involved in the analysis and interpretation of the results. SN, NSY, YGW, AM, and AH wrote the first draft of the manuscript. RI, ZH, FJ, and IN reviewed and provided scientific revisions to the manuscript. All the authors have read and approved the final manuscript. RI is responsible for the overall content as guarantor.

## Data availability statement

The datasets used and/or analyzed during the current study are available from the corresponding author upon reasonable request.

## Ethical Consideration

The study was approved by the Ethical Review Committee of the Aga Khan University (Ref 2022-5920-22763). Written informed consent was obtained from study participants before participation.

## Consent for Publication

Not applicable

## Acknowledgments

We thank all study participants who took part in this study. We are thankful to all frontline workers who provided services at PHCs in the community.

## Supplementary materials

**Table S1.** Equations for calculating composite risk thresholds from STRiDE-India Cohort.

|  | Composite risk score equations |
| --- | --- |
| **Model 1a** | only HbA1c based on their cut-offs |
| **Model 1b** | $-2.82813 + 0.041514 \cdot \text{age} + 0.005341 \cdot \text{BMI} + 0.3193 \cdot \text{FH DM}$ |
| **Model 2** | $-5.213 + 0.518 \cdot \text{HbA1C} + 0.037 \cdot \text{age} + 0.0003 \cdot \text{BMI} + 0.303 \cdot \text{FH DM}$ |
| **Model 3** | $-5.886 + 0.393 \cdot \text{HbA1C} + 0.040 \cdot \text{age} - 0.00036 \cdot \text{BMI} + 0.353 \cdot \text{FH DM} - 0.487 \cdot \text{SES Medium} - 0.351 \cdot \text{SES Upper} + 0.006 \cdot \text{GA} - 0.105 \cdot \text{Parity} + 0.010 \cdot \text{SBP} + 0.005 \cdot \text{DBP}$ |

**BMI**=body mass index, **FH DM**= family history of diabetes mellitus, **SES**=socioeconomic status, **GA**= gestational age, **SBP**= systolic blood pressure, **DBP**=diastolic blood pressure

**Table S2.** Baseline characteristics of the three risk groups according to the ‘rule-in’ and ‘rule-out’ approach: Model 2 (N=2485)

| Variables | Risk categories based on the 'rule-in' and rule-out' approach |  |  | P-value |
| --- | --- | --- | --- | --- |
|  | Rule out GDM<br>(lowest risk)<br>(n=1010) | Requires OGTT<br>(medium risk)<br>(n=1039) | Rule in to have GDM<br>(highest risk)<br>(n=436) |  |
| | mean ± SD or n (%) | mean ± SD or n (%) | mean ± SD or n (%) | |
| Age, years | 22.5 ± 3.3 | 27.5 ± 4.4 | 34.4 ± 5.0 | <0.001 |
| Height, cm | 153.7 ± 5.3 | 154.3 ± 5.6 | 154.7 ± 6.5 | 0.003 |
| Weight, kg | 49.0 ± 10.0 | 53.5 ± 12.1 | 59.3 ± 13.8 | <0.001 |
| BMI, kg/m<sup>2</sup> | 20.7 ± 3.9 | 22.4 ± 4.8 | 24.8 ± 5.6 | <0.001 |
| BMI categories, kg/m<sup>2</sup> |  |  |  |  |
| Underweight (<18.5) | 324 (32.1) | 229 (22.1) | 53 (12.1) |  |
| Normal weight (18.5-22.9) | 447 (44.3) | 392 (37.7) | 126 (28.9) |  |
| Overweight (23.0-26.9) | 158 (15.6) | 237 (22.8) | 118 (27.1) |  |
| Obesity (≥27.0) | 81 (8.0) | 181 (17.4) | 139 (31.9) | |
| Parity |  |  |  |  |
| Nulliparous | 378 (37.4) | 164 (15.8) | 28 (6.4) | <0.001 |
| Primiparous | 291 (28.8) | 213 (20.5) | 33 (7.6) |  |
| Multiparous | 341 (33.8) | 662 (63.7) | 375 (86.0) |  |
| Family history of diabetes | 21 (2.1) | 142 (13.7) | 149 (34.2) | <0.001 |
| SBP, mmHg | 101.9 ± 10.2 | 103.1 ± 11.2 | 106.5 ± 11.2 | <0.001 |
| DBP, mmHg | 62.1 ± 7.7 | 63.5 ± 8.1 | 66.1 ± 8.1 | <0.001 |
| GA at recruitment, weeks | 13.1 ± 3.5 | 12.9 ± 3.5 | 13.1 ± 3.3 | 0.40 |
| GA at OGTT, weeks | 28.1 ± 1.2 | 28.1 ± 2.1 | 28.0 ± 1.3 | 0.29 |
| HbA1c, % | 4.9 ± 0.3 | 5.2 ± 0.3 | 5.4 ± 0.3 | <0.001 |
| OGTT fasting glucose (mg/dL) | 73.5 ± 7.0 | 74.5 ± 7.2 | 76.3 ± 8.5 | <0.001 |
| OGTT 1 hour glucose (mg/dL) | 118.2 ± 24.6 | 125.4 ± 26.1 | 134.1 ± 29.7 | <0.001 |
| OGTT 2 hours glucose (mg/dL) | 101.4 ± 21.6 | 106.2 ± 23.5 | 112.6 ± 29.0 | <0.001 |

**Table S3.** Baseline characteristics of the three risk groups according to the ‘rule-in’ and ‘rule-out’ approach: Model 3 (N=2485)

| Variables | Risk categories based on the 'rule-in' and rule-out' approach |  |  | P-value |
| --- | --- | --- | --- | --- |
|  | Rule out GDM<br>(lowest risk)<br>(n=1377) | Requires OGTT<br>(medium risk)<br>(n=881) | Rule in to have GDM<br>(highest risk)<br>(n=227) |  |
|  | mean ± SD or n (%) | mean ± SD or n (%) | mean ± SD or n (%) |  |
| Age, years | 24.1 ± 4.2 | 28.8 ± 5.5 | 34.0 ± 6.3 | <0.001 |
| Height, cm | 153.9 ± 5.4 | 154.4 ± 6.0 | 154.1 ± 6.1 | 0.12 |
| Weight, kg | 50.3 ± 10.7 | 54.8 ± 12.9 | 58.9 ± 14.0 | <0.001 |
| BMI, kg/m<sup>2</sup> | 21.2 ± 4.2 | 22.9 ± 5.1 | 24.8 ± 5.8 | <0.001 |
| BMI categories, kg/m<sup>2</sup> |  |  |  |  |
| Underweight (<18.5) | 407 (29.6) | 169 (19.2) | 30 (13.2) |  |
| Normal weight (18.5-22.9) | 576 (41.8) | 323 (36.7) | 66 (29.1) |  |
| Overweight (23.0-26.9) | 248 (18.0) | 211 (23.9) | 54 (23.8) |  |
| Obesity (≥27.0) | 146 (10.6) | 178 (20.2) | 77 (33.9) |  |
| Parity |  |  |  |  |
| Nulliparous | 376 (27.3) | 170 (19.3) | 24 (10.6) | <0.001 |
| Primiparous | 345 (25.1) | 161 (18.3) | 31 (13.7) |  |
| Multiparous | 656 (47.6) | 550 (62.4) | 172 (75.8) |  |
| Family history of diabetes | 56 (4.1) | 176 (20.0) | 80 (35.2) | <0.001 |
| SBP, mmHg | 100.0 ± 9.6 | 105.8 ± 10.3 | 112.0 ± 12.6 | <0.001 |
| DBP, mmHg | 61.2 ± 7.2 | 65.3 ± 7.6 | 69.4 ± 9.6 | <0.001 |
| GA at recruitment, weeks | 12.8 ± 3.5 | 13.2 ± 3.4 | 13.3 ± 3.5 | 0.004 |
| GA at OGTT, weeks | 28.0 ± 1.3 | 28.2 ± 2.2 | 28.1 ± 1.3 | 0.07 |
| HbA1c, % | 5.0 ± 0.3 | 5.2 ± 0.3 | 5.4 ± 0.3 | <0.001 |
| OGTT fasting glucose (mg/dL) | 73.5 ± 6.8 | 75.3 ± 8.0 | 76.8 ± 7.5 | <0.001 |
| OGTT 1 hour glucose (mg/dL) | 120.3 ± 24.9 | 127.3 ± 27.6 | 133.7 ± 30.0 | <0.001 |
| OGTT 2 hours glucose<br>(mg/dL) | 102.7 ± 22.5 | 107.8 ± 24.7 | 111.5 ± 29.1 | <0.001 |

